# Effects of an innovative daily physical activity intervention on the health and fitness status of primary school children: a cluster randomized controlled trial

**DOI:** 10.1101/2023.02.22.23286304

**Authors:** Gerald Jarnig, Reinhold Kerbl, Mireille N.M. van Poppel

**Affiliations:** Institute of Human Movement Science, Sport and Health, University of Graz, 8010 Graz, Austria; Department of Pediatrics and Adolescent Medicine, LKH Hochsteiermark, 8700 Leoben, Austria

## Abstract

**Aims:** An important barrier for a nationwide implementation of a daily physical activity (PA) intervention at primary schools is the lack of spatial and human resources. Therefore, the aim was to develop an innovative PA intervention that can be implemented, without additional spatial and human resource requirements. The effects of this intervention on anthropometrics and health-related fitness of primary school children were investigated.

**Methods:** In twelve primary schools, 24 classes were randomly allocated to an intervention and control group. In the intervention group, children received mix physical education lessons, cognitive content using movement, and additional opportunities for extracurricular physical activities (home exercises) over a 9-month period. In the control group, children received the usual physical education classes. At baseline (Sept 2021) and at follow-up immediately after the intervention (June 2022), body weight, height, waist circumference and fitness parameters (cardiorespiratory fitness, muscle strength, flexibility) were measured. Intervention effects were assessed using multilevel regression models, adjusted for gender, urban/rural school, sports club membership, and for baseline values of the outcome parameter. Interactions between intervention and being a member of a sports club were assessed.

**Results:** Of 485 invited children, 412 (85%) were included in the analyses; 228 in the intervention and 184 in the control group. Children were 9.7 (SD 0.5) years old at baseline. In the total intervention group, a reduction in waist-to-height ratio and increase in all fitness parameters were found compared to the control group. Interactions with being a member of a sports club were found, and intervention effects were more pronounced in the group of children who were not a member of a sports club.

**Conclusions:** Our daily physical activity intervention for primary schools showed positive effects on important health related parameters, especially for children who were not in a sports club and had most of their PA at school. Wider implementation of this intervention in primary schools is warranted, which should be feasible, since we made sure that no extra resources are needed.

**Trial registration:** DRKS00025515.

## Introduction

Adequate physical activity in childhood and adolescence leads to short-term and lifelong health benefits [1–4] and has additional positive effects on cognitive performances [5,6]. Especially cardiorespiratory fitness (CRF) and muscular fitness (MF) in childhood and adolescence are important health markers [7], which are associated with lower body mass index (BMI), smaller waist circumference, reduced body fat percentage, and lower prevalence of metabolic syndrome later in life [8].

Overall fitness improves with higher levels of physical activity (PA) [9–11]. Daily PA in childhood and adolescence is the sum of PA performed during transportation (walking, cycling, etc.) to and from school, while at school, participating in sports club training, and through active play in leisure time [12,13]. In recent years, the participation in organized sports slightly increased, however in contrast, the percentage of children who participated in unorganized sports and outdoor play in leisure time decreased [14,15]. In general, just a small proportion (29%) of European children and adolescents achieve the daily PA recommendation issued by the world health organization (WHO), and more than two-thirds of children are insufficiently active [16]. Generally, children who are member of a sports club are more often achieve the health enhancing physical activity recommendation [17,18] of 60 minutes of at least moderate PA daily [19] compared to children who are not member of a sports club.

The COVID-19 pandemic dramatically intensified this worrisome situation, of lack of adequate PA on a daily basis, as children and adolescents’ total daily PA decreased by 20% during the COVID-19 pandemic, compared to pre-Covid-19 level [20].

Worldwide, different strategies are developed to increase PA in the overall population [21]. For school-aged children, the main focus is to implement interventions at school [22–28], and a daily PA program is a frequently used measure [29], which has already been tested and proven to be very effective for improving PA levels in many regional projects [30–32].

However, for a nationwide implementation of the daily PA program in schools, limited structural resources, such as sports halls and teachers or sports coaches, in the primary school environment pose a major problem [33,34].

To enable a nationwide implementation of a daily PA program at primary schools without additional spatial and human resources, a completely innovative concept for a daily physical activity session was developed, and the effects of this program on BMI and health-related fitness of primary school children were evaluated.

## Materials and Methods

### Design

The study was designed as a parallel, cluster randomized controlled trial (intervention group vs. control group, 1:1 ratio) to evaluate the effects of a daily PA program on BMI and health-related fitness of primary school children. The study was carried out in the greater Klagenfurt area, Austria, between September 2021 and June 2022, and was registered in the German Clinical Trials Registry (ID DRKS00025515) and approved by the Research Ethics Committee of the University of Graz, Styria, Austria (GZ. 39/23/63 ex 2018/19).

### Selection of schools and participants

A list of all 39 primary schools in the urban and rural districts of Klagenfurt, Austria, was used to select the schools. A random number generator was used to select 12 schools, stratified by rural and urban districts. All administrators of the selected schools consented to the participation of their school classes in the study. In spring 2021, all 485 children attending third grade at that time in one of the 12 schools were invited to participate in the study. A total of 467 (96.3%) legal guardians gave their written consent for the participation of their children. Inclusion criteria were that the children participating in the study had no physical limitations, were aged between 8 and 12 years at baseline, and were attending fourth grade in September 2021. One child was excluded from study participation because it did not meet all inclusion criteria. Additionally, children who were absent from school on more than 25% of school days between September 2021 and June 2022 were excluded from the study analysis.

### Randomization and blinding

Using a random number generator, the 24 fourth grade school classes, that were part of one of the previously selected 12 schools, were allocated to an intervention group (IG) and a control group (CG), stratified by district (Figure 1). Baseline assessments were carried out blinded for allocation, further blinding of participants or assessments was not possible.

**Figure 1.**
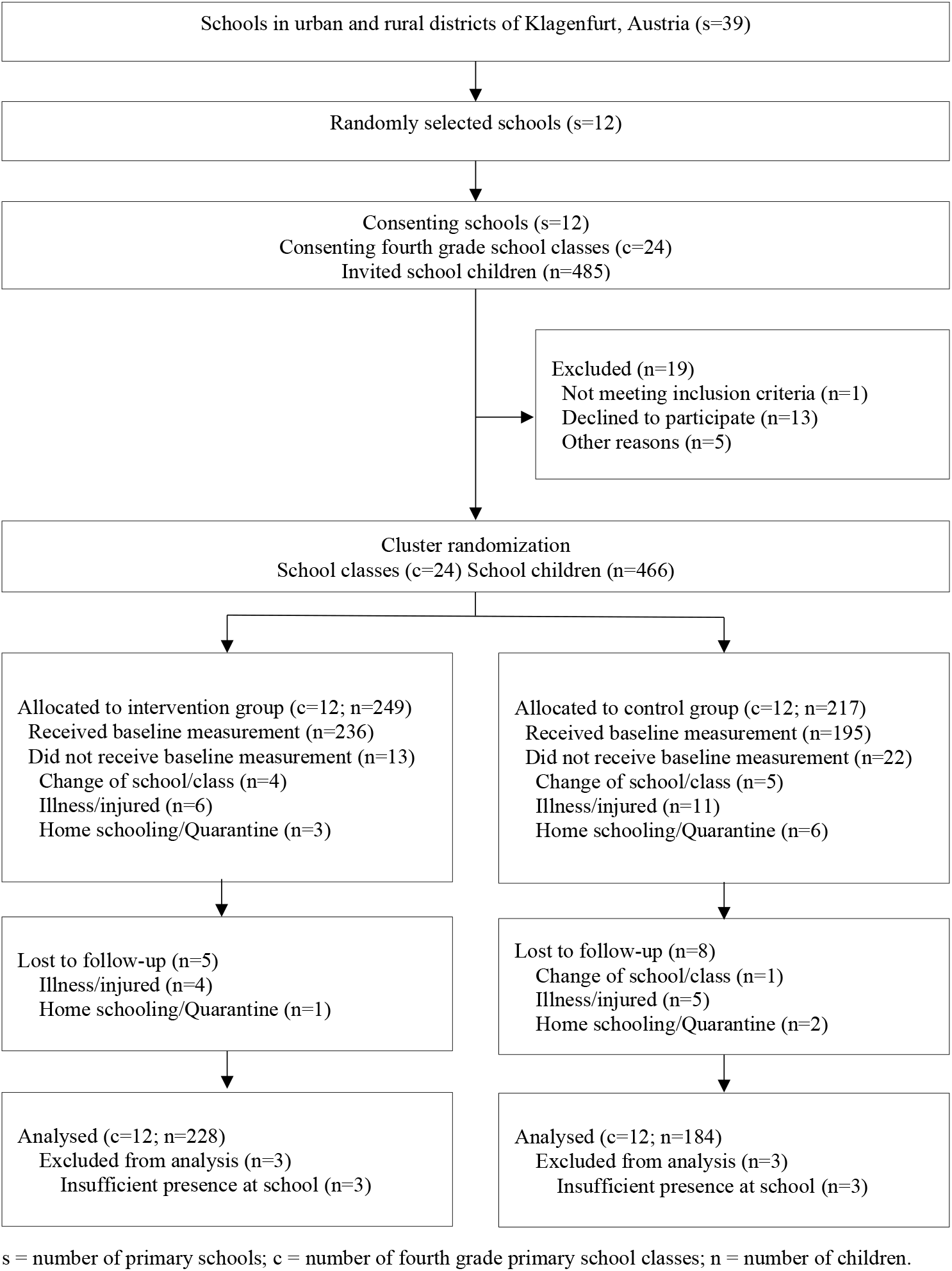
Flow diagram.

### Intervention program

The aim of the intervention was to achieve a daily PA lesson in school. A daily school lesson in the IG, which in Austria at primary school lasts 50 minutes, was carried out with adequate PA during and after school hours in order to achieve this goal.

The main focus in planning the intervention program was that, in case of a positive evaluation of the resulting effects, it could be implemented nationwide as quickly as possible without additional necessary spatial or personnel resources and without changing the school curriculum. For this reason, all elements of the intervention were planned in advance by external experts and documented on more than 800 pages in written and illustrated form in order to enable optimal and efficient nationwide implementation at a later date.

The PA intervention program consisted of two main modules (I&II) and one additional module (III):

I) The physical education (PE) classes of the children in the IG were planned by external experts, according to the Austrian school curriculum. In this curriculum, PE classes are scheduled twice per week for 50 minutes. In the intervention group, the PE classes in the schools were provided not by the usual teacher, but by external coaches, who were specifically trained and who weekly received the planned content for the PE classes.

II) On the three days per week that there were no PE classes, the children learned cognitive content combined with specific movement activities (in mathematics and language art) in one school hour of 50 minutes. For the planning of these specific PA-supporting lessons, work materials were developed by external primary school teachers in collaboration with sports experts, and these materials were provided to all class teachers and implemented by them on site in the school.

III) PA activities were given as homework once a week. For example, the children were instructed to go for a walk in the countryside and were given a checklist of objects to be seen on this walk, such as a red car, a park bench, or a pedestrian crossing. If an object was spotted, the children had the ticking it off on the checklist; and when all objects were seen, the mission was successfully completed. The work materials for this PA-supporting homework were developed by external sports experts, and provided to all class teachers, who handed out the materials to the children.

The intervention program was started after the baseline measurement (T1) in September 2021 and was implemented for nine months. The control group received the usual school curriculum, consisting of PE classes twice a week for 50 minutes.

### Outcome measurements

All measurements were standardized and pilot tested with trained personnel (sports scientists) with non-participating children (n = 98) of the same age group before the study began. Baseline measurements (T1) were conducted in September 2021, and after intervention ended, the second test phase (T2) was carried out in mid-June 2022. All anthropometric data and fitness tests were collected barefoot and in sportswear. Baseline data (T1) were collected before randomization by four sports scientists, who were blinded for intervention allocation. Two of them also led the subsequent physical activity sessions in the intervention classes, which took place in the gym or on the sports field as part of curriculum. The follow-up measurement (T2) was carried out by the same testing team. The anthropometrics data, as well as the data for 6-minute run (6MR), shuttle run test (4 × 10 SHR) and jumping sideways (JS) were collected by the two sports scientists not involved in the intervention, and who were thus still blinded for the allocation to IG or CG. However, data for standing long jump (SLJ), two-kilogram medicine ball throw (MB2kg), push-up test (PU), and V-Sit-and-Reach-test (VSR) were collected by the two sports scientists who were not blinded any longer. The tests were completed in one day, with the 6MR performed at the end.

### Primary and secondary outcomes

To assess the intervention’s impact on health status, body mass index (BMI) [35,36] and waist-to-height ratio (WtHR) [37–39] were primary outcomes of this study. In addition, health-related fitness (HRF) was a primary outcome, consisting of cardiorespiratory endurance, muscular strength, and flexibility of primary school children.

As secondary outcome, action speed, a parameter of performance-related fitness, was measured.

### Anthropometrics data

Weight (kg) was measured to the nearest 0.1 kg using a Bosch PPW4202/01 body scale. Height (cm) was measured to the nearest 0.1 cm using a SECA 213 stadiometer, and waist circumference (cm) of the children was measured to the nearest 0.1 cm using a GIMA 27343 body tape measure.

The crude BMI (body weight in kg divided by height squared in meters) and the waist-to-height ratio (WHtR – waist circumference in cm divided by height in cm) were calculated.

### Health related fitness

HRF includes cardiorespiratory endurance, muscular strength, and flexibility [9]. Thus, the 6-minute run (6MR; for cardiorespiratory endurance), standing long jump (SLJ; for lower body muscular strength), two-kilogram medicine ball throw (MB2kg; for upper body muscular strength), jumping sideway (JS; for lower body muscular endurance), push-up test (PU; for upper body muscular endurance), and sit-and-reach test (VSR; for flexibility) were used. All used fitness tests are internationally established, frequently used, and have very good test quality criteria (objectivity, reliability, validity) [40,41].

#### 6-minute run (6MR)

For 6 minutes, the children were instructed to run continuously around a rectangle (6 m × 18 m) and to run a maximum possible distance in this time. The test was completed by seven children simultaneously, there was one scoring attempt and the running performance was measured to the nearest meter.

#### Standing long jump (SLJ)

From a starting line, the children had to jump with both legs as far as possible. The minimum distance between the starting line and the contact of the child’s heels with the ground was measured with a tape measure to the nearest cm. The children had three scoring attempts, and only the best score was used.

#### Two-kilogram medicine ball throw (MB2kg)

From a starting line, the children had to throw a 2 kg medicine ball, which was held with both hands and touched the chest, as far forward as possible. The minimum distance between the starting line and the ball’s contact with the ground was measured to the nearest centimeter with a tape measure. The best of two scoring attempts was used.

#### Jumping sideway (JS)

Children were instructed to jump sideways over a wooden stick (60 cm 4 cm 2 cm) as many times as possible in 15 seconds. The jumps had to be performed using both feet at the same time and the children had two scoring attempts. The sum total of the number of jumps from both attempts was used.

#### Push-up test (PU)

Starting position: The child was lying in the prone position on the floor and the hands touched on the buttocks. After the start command, the hand contact was released, the hands touched the floor next to the shoulders and pushed the body up into a push-up position as straight extended as possible. Then the child released one hand from the floor and touched the other hand with it. The rest of the body remained straight extended, only the hands and feet were in contact with the floor. The hand was returned to the position on the floor and the elbows were bent until the body was again in prone position. The hands touched the buttocks again in the prone position, then a push-up was completed. The knees did not make contact with the ground during the whole test. The children had 40 seconds to complete as many correctly performed push-ups as possible. One scoring attempt was made and the number of correctly performed push-ups was used.

#### Flexibility (VSR)

The V-Sit-and-Reach-Test (VSR) [41] was selected to measure flexibility, a simple variation of the classic Sit-and-Reach-Test being carried out without additional costs. A tape measure and marking tape were needed to perform it. A heel line was marked with the tape and the tape measure was fixed to the floor. The children sat down on the floor with their legs spread 30-40 cm apart, their heels were placed on the heel line, then the children placed one hand on top of the other and slowly stretched forward as far as possible. The distance between the heel line and the maximum reached position, which could be held with the fingertips for two seconds, was noted. Each child had two scoring attempts; the longest distance reached was used. To be able to use reference values from the classic Sit and Stretch test, 15 cm was added to the scoring attempt.

### Secondary outcome

#### Action Speed (4 × 10 SHR)

A shuttle run test (4 × 10 SHR) was performed for assessing the children’s action speed. A starting and turning line was marked on the floor at a distance of 10 m using marking tape. Three easy-to-grasp pieces of foam (S1, S2, S3) were needed (about 5.0 × 5.0 × 3.0 cm). S1 and S3 were placed behind the turning line and S2 in front of the starting line. Children had to run from the starting line crossing the turning line, pick up S1, run back crossing the starting line, and put S1 down. Then they picked up S2, ran again crossing the turning line, put down S2, picked up S3, and ran with it crossing the starting line. The children were instructed to run as fast as possible completing this test. Two scoring attempts were made, and the time was measured with a stopwatch to the nearest 0.01 seconds. Each child had two attempts, and the fastest run was used.

### Sample size

We calculated with G*Power that a total sample size of 423 children was required for detecting an effect size d of 0.30, with a power of 80% and alpha of 5% in a two-tailed test, and assuming a 20% drop out (more details are provided in the supplementary materials).

### Moderation

Children with sports club memberships have a greater amount of average weekly PA outside of the school settings [14] and mostly show healthier/better health status/health-related fitness scores [42–46]. For these reasons, we assessed whether being a member of a sports club moderated the effects of the intervention. When completing the consent form, legal guardians provided in writing information whether or not their child had a sports club membership at the baseline measurement.

### Standardization

#### Anthropometric data

Standard deviation scores (SDS) for BMI and WtHR were calculated using the LMS method [47] based on recent international age- and sex-specific references. To calculate SDS for BMI, we used the internationally used reference values of the International Obesity Taskforce (BMI_IOTF_ SDS) [48].

Since there are no national reference values of the WtHR, we used international reference values from Poland from 2016 for the SDS calculation (WtHR SDS) [49]. Whereas healthier and lower raw values of WtHR were expressed with higher positive SDS and, in contrast, unhealthier and higher raw values of WtHR were described with higher negative SDS.

#### Fitness parameters

To compare the results (raw score) of the fitness tests with established reference values, standard deviation scores (SDS) and traditional z-scores (z-values) were created based on age- and sex-specific reference values. Since no Austrian national reference values were available for this age group, international reference values were used. For the 6MR, the SLJ, and the MB2kg, the most recent German percentile tables of the Düsseldorf Model ([50]; collected 2011-2018) were used, for JS and 4 × 10 SHR, Portuguese norm values [51] from the Motor Competence Assessment (MCA) of 2019 were used, and for VSR, Greek [52] standard values from the year 2015 were used. For these international references, calculations were performed based on the LMS method [47] using the German (DüMo), Portuguese (MCA), and Greek (GRE) reference tables. For the PU, German normative values [53] of the German Motor Test (GMT) from 2016 were used and z values were calculated by a traditional z-value standardization.

### Statistical analysis

Continuous variables are reported as means (M) and standard deviations (SD) and categorical variables as absolute values (n) and percentages (%) for descriptive statistics. No imputation of the data was performed.

To determine effects of the intervention on health status (BMI_IOTF_ SDS, WHtR) and parameters of fitness (6MR SDS, SLJ SDS, MB2kg SDS, JS SDS, PU z-value, VSR SDS and 4 × 10 SHR SDS), multilevel analyses were undertaken with a two-level structure: individual and school. The SDS values at follow-up were dependent variables, and intervention group (IG=1 and CG=0) the independent variable. Models were adjusted for gender (girls=1 and boys=0), school location (urban=1 and rural=0), sports club membership (no sports club=1 and sports club=0) and respective baseline SDS values. To assess possible effect moderation of being a member of a sports club, the interaction term (intervention group*sports membership) was added. Since interactions were present (p<0.10 for the interaction term), analyses were performed for the groups of children with and without a sports club membership separately.

All tests were two-tailed, with a *p*-value < 0.05 considered statistically significant. All statistical calculations were performed using SPSS Version 28 (IBM Corp. Released 2021. IBM SPSS Statistics for Windows, Armonk, NY: IBM Corp).

## Results

In September 2021, 431 children participated in the baseline measurements (T1). Thirteen children did not participate in the follow-up measurement in June 2022 (T2) and six children were present at school on less than 75% of all school days between T1 and T2. These 19 children were excluded from analyses, resulting in 412 children (95.6% from baseline) with complete data (Figure 1).

Of the 412 children, 228 (55.3%) participated in the intervention. Baseline characteristics of the study population are described in Table 1. Children had a mean (SD) age of 9.7 (0.5) years, 196 (47.6%) were girls, 171 (41.5%) were members of a sports club, and 257 (62.4%) lived in the urban region of Klagenfurt (Table 1). Differences in baseline values between intervention (IG) and control group (CG) were found for the outcomes JS SDS (*p* = 0.005) and VSR SDS (*p* = 0.033) (Table 2)

**Table 1.** Baseline characteristics of participants included in the analyses, by intervention group.

| Variable | Total<br>*n* =412 | Intervention<br>*n* =228 | Control<br>*n* =184 | *P* value |
| --- | --- | --- | --- | --- |
| Urban School, No. (%) | 257 (62.4%) | 147 (64.5%) | 110 (59.8%) | 0.33 |
| Age (years), mean (SD) | 9.7 (0.5) | 9.7 (0.5) | 9.8 (0.5) | 0.19 |
| Girls, No. (%) | 196 (47.6%) | 103 (45.2%) | 93 (50.5%) | 0.28 |
| Member of sports club, No. (%) | 171 (41.5%) | 101 (44.3%) | 70 (38.0%) | 0.20 |

Data are the No. (%) or mean (SD). SD = standard deviation, *P* value = in bold indicate statistical significance ( $p < 0.05$ ).

**Table 2.**
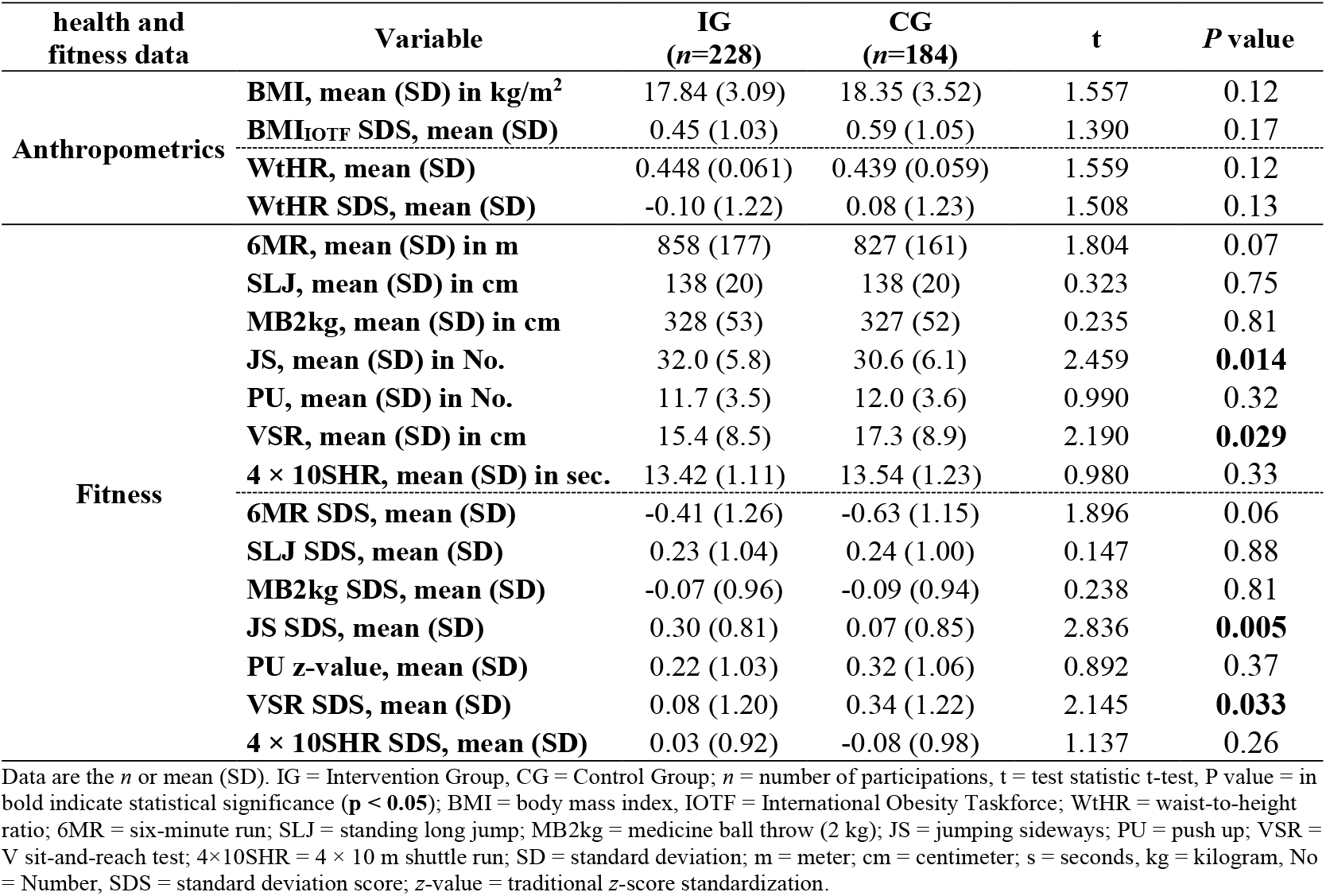
Differences in raw fitness data and standard deviation scores (SDS) at baseline by intervention group (intervention vs control).

| health and fitness data | Variable | IG<br>(*n*=228) | CG<br>(*n*=184) | *t* | *P* value |
| --- | --- | --- | --- | --- | --- |
| Anthropometrics | BMI, mean (SD) in kg/m<sup>2</sup> | 17.84 (3.09) | 18.35 (3.52) | 1.557 | 0.12 |
|  | BMI<sub>IOTF</sub> SDS, mean (SD) | 0.45 (1.03) | 0.59 (1.05) | 1.390 | 0.17 |
|  | WtHR, mean (SD) | 0.448 (0.061) | 0.439 (0.059) | 1.559 | 0.12 |
|  | WtHR SDS, mean (SD) | -0.10 (1.22) | 0.08 (1.23) | 1.508 | 0.13 |
| Fitness | 6MR, mean (SD) in m | 858 (177) | 827 (161) | 1.804 | 0.07 |
|  | SLJ, mean (SD) in cm | 138 (20) | 138 (20) | 0.323 | 0.75 |
|  | MB2kg, mean (SD) in cm | 328 (53) | 327 (52) | 0.235 | 0.81 |
|  | JS, mean (SD) in No. | 32.0 (5.8) | 30.6 (6.1) | 2.459 | **0.014** |
|  | PU, mean (SD) in No. | 11.7 (3.5) | 12.0 (3.6) | 0.990 | 0.32 |
|  | VSR, mean (SD) in cm | 15.4 (8.5) | 17.3 (8.9) | 2.190 | **0.029** |
|  | 4 × 10SHR, mean (SD) in sec. | 13.42 (1.11) | 13.54 (1.23) | 0.980 | 0.33 |
|  | 6MR SDS, mean (SD) | -0.41 (1.26) | -0.63 (1.15) | 1.896 | 0.06 |
|  | SLJ SDS, mean (SD) | 0.23 (1.04) | 0.24 (1.00) | 0.147 | 0.88 |
|  | MB2kg SDS, mean (SD) | -0.07 (0.96) | -0.09 (0.94) | 0.238 | 0.81 |
|  | JS SDS, mean (SD) | 0.30 (0.81) | 0.07 (0.85) | 2.836 | **0.005** |
|  | PU z-value, mean (SD) | 0.22 (1.03) | 0.32 (1.06) | 0.892 | 0.37 |
|  | VSR SDS, mean (SD) | 0.08 (1.20) | 0.34 (1.22) | 2.145 | **0.033** |
|  | 4 × 10SHR SDS, mean (SD) | 0.03 (0.92) | -0.08 (0.98) | 1.137 | 0.26 |

Data are the *n* or mean (SD). IG = Intervention Group, CG = Control Group; *n* = number of participations, *t* = test statistic t-test, *P* value = in bold indicate statistical significance ( $p < 0.05$ ); BMI = body mass index, IOTF = International Obesity Taskforce; WtHR = waist-to-height ratio; 6MR = six-minute run; SLJ = standing long jump; MB2kg = medicine ball throw (2 kg); JS = jumping sideways; PU = push up; VSR = V sit-and-reach test; 4×10SHR = 4 × 10 m shuttle run; SD = standard deviation; m = meter; cm = centimeter; s = seconds, kg = kilogram, No = Number, SDS = standard deviation score; z-value = traditional z-score standardization.

### Primary outcomes

In the supplements, the raw and standardized scores of all outcomes at baseline and after the intervention are provided (Table S1 & S2). We found a highly statistically significant improvement in WtHR SDS (unstandardized regression coefficient (B) = 0.30, *p* < 0.001) in the IG compared to the CG. In contrast, no statistically significant changes were observed between the IG and CG groups in the development of BMI_IOTF_ SDS (B = 0.01, *p* = 0.68) (Table 3, Figure 2).

**Table 3.** Multilevel regression assessing the association of the group membership (intervention or control) on the development of health status and fitness parameters, for all and for subgroups sports club membership.

| *health and fitness status* | All<br>I vs C<br>(*n*=412) |  | No Sports Club<br>I vs C<br>(*n*=241) |  | Sports Club<br>I vs C<br>(*n*=171) |  |
| --- | --- | --- | --- | --- | --- | --- |
|  | B (95% CI) | *P* value | B (95% CI) | *P* value | B (95% CI) | *P* value |
| BMI<sub>IOTF</sub> SDS | 0.01 (-0.05; 0.07) | 0.68 | 0.02 (-0.07; 0.10) | 0.67 | 0.00 (-0.09; 0.09) | 0.99 |
| WtHR SDS | 0.30 (0.16; 0.44) | **< 0.001** | 0.34 (0.16; 0.53) | **< 0.001** | 0.25 (0.04; 0.47) | **0.020** |
| 6MR SDS | 0.20 (0.01; 0.38) | **0.037** | 0.38 (0.16; 0.61) | **< 0.001** | -0.07 (-0.38; 0.24) | 0.64 |
| SLJ SDS | 0.11 (0.00; 0.21) | **0.041** | 0.16 (0.03; 0.30) | **0.025** | 0.03 (-0.13; 0.19) | 0.74 |
| MB2kg SDS | -0.06 (-0.20; 0.07) | 0.36 | 0.01 (-0.18; 0.19) | 0.95 | -0.18 (-0.39; 0.03) | 0.10 |
| JS SDS | 0.12 (0.01; 0.23) | **0.027** | 0.19 (0.04; 0.34) | **0.015** | 0.03 (-0.13; 0.18) | 0.73 |
| PU z-value | -0.12 (-0.27; 0.04) | 0.13 | -0.06 (-0.27; 0.15) | 0.58 | -0.19 (-0.41; 0.03) | 0.08 |
| VSR SDS | 0.19 (0.03; 0.35) | **0.019** | 0.31 (0.10; 0.53) | **0.005** | 0.00 (-0.22; 0.23) | 0.99 |
| 4 × 10SHR, SDS | 0.10 (-0.03; 0.22) | 0.12 | 0.20 (0.04; 0.36) | **0.017** | -0.05 (-0.24; 0.14) | 0.60 |

Adjusted for baseline, gender, school location, sports club membership (total only); I = Intervention group, C = Control group, *n* = number of participations, B = unstandardized regression coefficient, CI = Confidence Interval, *P* value = in bold indicate statistical significance (***p* < 0.05**); BMI = body mass index, IOTF = International Obesity Taskforce; WtHR = waist-to-height ratio; 6MR = six-minute run; SLJ = standing long jump; MB2kg = medicine ball throw (2 kg); JS = jumping sideways; PU = push up; VSR = V sit-and-reach test; 4×10SHR = 4 × 10 m shuttle run; SDS = standard deviation score; z-value = traditional z-score standardization;

**Figure 2.**
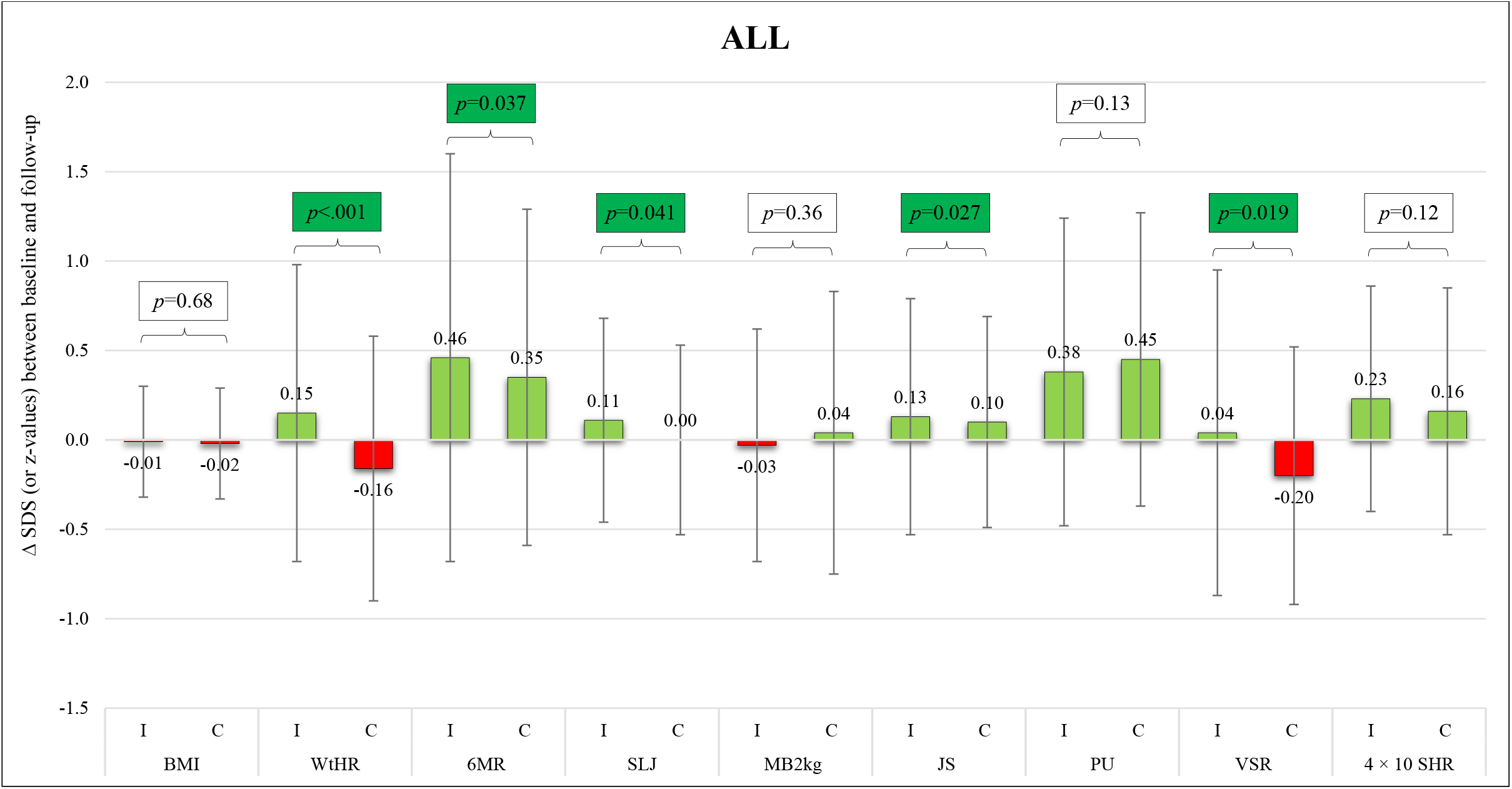
Changes in standard deviation scores between baseline and follow-up in the intervention and control groups

The results of the health-related fitness tests showed statistically significant improvements in the IG compared to the CG in cardiorespiratory endurance (6MR SDS: B = 0.20, *p* = 0.037), lower body muscle strength (SLJ SDS: B = 0.11, *p* = 0.041), lower body muscle endurance (JS SDS: B = 0.12, *p* = 0.027), and flexibility (VSR SDS: B = 0.19, *p* = 0.019) (Table 3, Figure 2). No statistically significant differences between IG and CG were detected in upper body muscle strength (MB2kg SDS: B = −0.06, *p* = 0.36), upper body muscle endurance (PU SDS: B = −0.12, *p* = 0.13) and action speed (4 × 10 SHR SDS: B = 0.10, *p* = 0.12) (Table 3, Figure 2).

### Secondary outcomes

No statistically significant differences between IG and CG were detected in action speed (4 × 10 SHR SDS: B = 0.10, *p* = 0.12) (Table 3, Figure 2).

### Moderation by sports club membership

Since relevant interaction was present, we performed separate analyses for the subgroups of children without a sports club membership (n=241) and children with a sports club membership (n=171).

Among children who were not a member of a sports club, highly statistically significant differences were found in the waist-to-height ratio (WtHR SDS: B = 0.34, p<0.001) and cardiorespiratory endurance (6MR SDS: B = 0.38, p <0.001) between the IG and CG (Table 3, Figure 3). Additionally, statistically significant differences were detected in lower body muscle strength (SLJ SDS: B = 0.16, p = 0.025), lower body muscle endurance (JS SDS: B = 0.19, p = 0.015), flexibility (VSR SDS: B = 0.31, p = 0.005), and action speed (4 × 10 SHR SDS: B = 0.20, p = 0.017) in IG compared to CG (Table 3, Figure 3). No significant differences were found between IG and CG in BMI (BMI SDS: B = 0.02, p = 0.67), upper body muscle strength (MB2kg SDS: B = 0.01, p = 0.95) or upper body muscle endurance (PU SDS: B = −0.06, p = 0.58) (Table 3, Figure 3).

**Figure 3.**
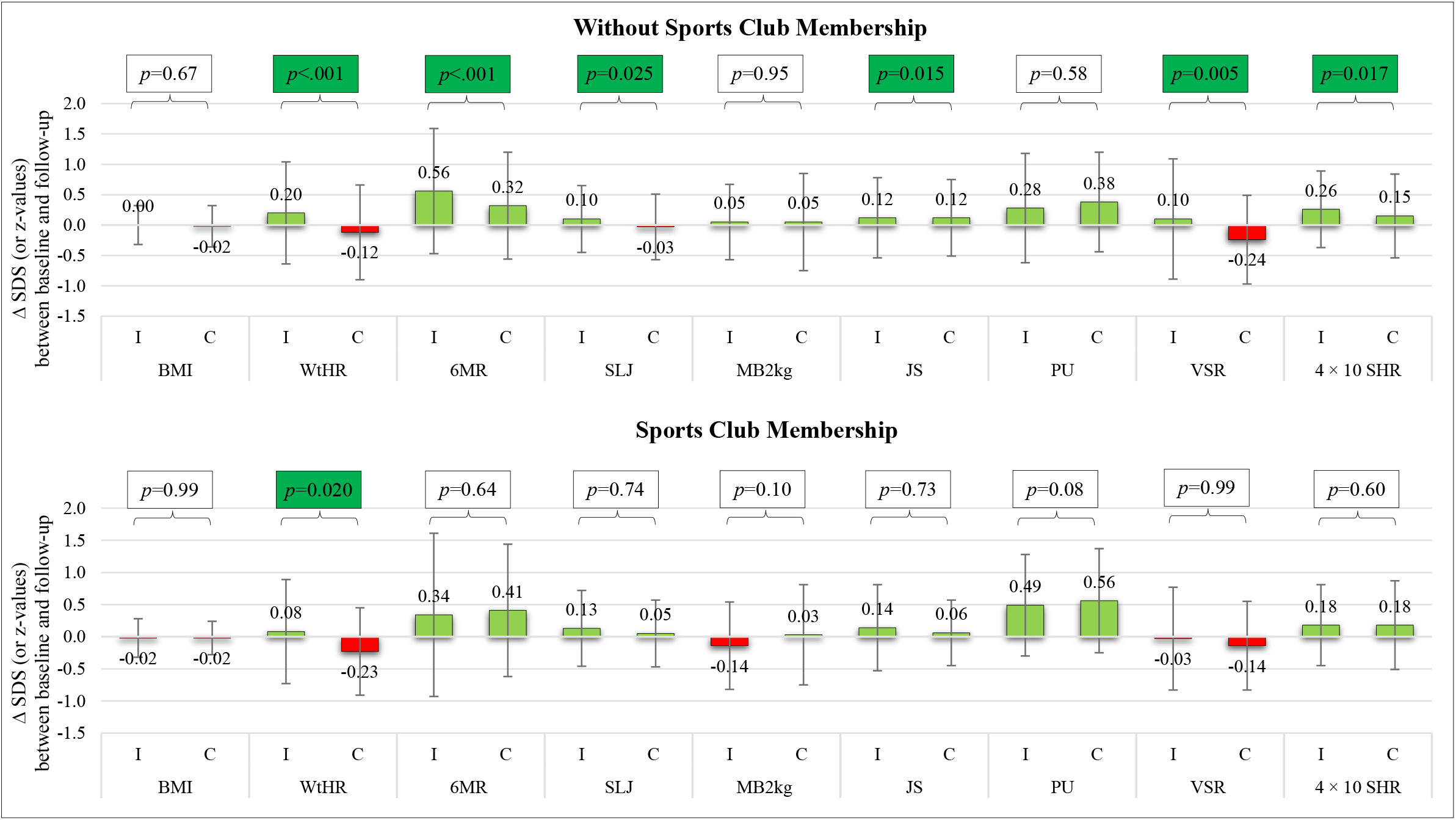
Changes in standard deviation scores between baseline and follow-up in the intervention and control groups, separately for children without and with sports club membership.

Among the children with sports membership, significant differences were found for waist-to-height ratio (WtHR SDS: B = 0.25, p = 0.02) in IG compared to CG, and no differences were found between IG and CG for BMI, health-related fitness parameters (6MR, SLJ, MB2kg, JS, PU, VSR), or action speed (4 × 10 SHR) (Table 3, Figure 3).

## Discussion

In the present study, we investigated the effects of a daily PA intervention program in primary schools on anthropometrics and health-related fitness. We found that in the intervention group, all health-related fitness parameters (cardiorespiratory endurance, muscle strength, flexibility) improved compared to the control group. These improvements were more pronounced in children who were not a member of sports club.

That our intervention was more effective for children who were not in a sports club is an important finding. It shows that fitness of these children is much more dependent on physical activity in the school setting. Therefore, especially for this group of children, providing a daily PA intervention is of utmost importance. That such an approach is less of importance for children who do sports outside of school, is to be expected, and is not a reason not to implement a PA intervention. They will benefit in other aspects, such as respect for rules, fair play or teamwork [54,55].

Our intervention goal was certainly not new; various different strategies have been used around the world to increase the amount of physical activity in the school settings [56] and different kinds of interventions have been used to achieve this goal [57]. In primary schools, physical education is often carried out by the class teacher, which results in a weakening of the priority given to this subject compared to other school subjects [58]. Therefore, the approach taken in many intervention programs is to use external sports coaches to teach the physical education classes [59]. Previously, the concept of teaching cognitive content in combination with movement and PA was used, in particular in mathematics and language learning, as in our intervention, and led to an improvement in the cognitive performance of the participating children [60,61]. Other intervention designs prefer to organize additional sports activities in the school settings outside of class time [59], adding an additional daily physical activity lesson to the curriculum [30,32,62], spending the daily morning break at school with physical activities [63], or implementing high-intensity physical activity programs during short breaks in cognitive lessons [64].

However, for a nationwide implementation of the above-mentioned concepts many barriers have to be overcome: either an external source of funding is necessary, additional time, spatial and human resources on the part of the school community would have to be provided or, as for example in the case of the PA breaks during the cognitive lessons, class teachers have to be convinced, since they see PA breaks as time-consuming and difficult to implement in practice [65].

In contrast, our concept does not require any changes in the school’s spatial or staffing structure, there are no additional costs in the day-to-day running of schools, and movement during lessons is an integral part of the cognitive content of math and language arts and therefore does not interrupt the flow of children’s learning.

### Strengths and limitations

The main strength is that our intervention can be implemented without additional lessons or spatial resources. This will improve feasibility of wider implementation enormously.

Furthermore, the program was evaluated in a randomized controlled trial, with measurement of anthropometric data and all fitness assessments performed by trained research assistants using validated measures.

A main strength is that all elements of the intervention were planned by external experts and documented on more than 800 pages in written and illustrated form, and thus a nationwide implementation in the fourth grade of primary school would be possible immediately without additional costs, since PE classes could also be carried out by the classroom teachers, using the planned content.

However, there are some limitations that should be noted. We did not use accelerometers or other devices to measure the daily physical activity of the study participants, and we did not have a measure for the intensity of the physical activity during the intervention. However, given the increase of fitness compared to the control group, the intervention as a whole was intensive and frequent enough to positively influence this important parameter. The success of combining cognitive learning content with movement and physical activity depended strongly on the motivation level of the teaching classroom teachers [66].

## Conclusion

Our daily physical activity intervention for primary schools showed positive effects on important health related parameters, especially for children who were not in a sports club and had most of their PA at school.

It might be preferable to have PE lessons in primary schools carried out by specifically trained PE teachers, since this would improve the quality of the lessons [67] and increase the perceived importance of physical activity and sport as a school subject [58], as is already the case in secondary schools. However, when this is not possible, the next best option is to have class teachers provide PE lessons based on a predefined curriculum, such as developed and evaluated in this study. Wider implementation of this intervention in primary schools is warranted, which should be feasible, since we made sure that no extra resources are needed.

## Supporting information

supplements Jarnig et al

## Data Availability

All data produced in the present study are available upon reasonable request to the authors

## Acknowledgments

This study was organized by the non-profit association NAMOA—Nachwuchsmodell Austria. The authors would like to thank all participants and their guardians; the trainers and staff of this study; Wolfgang Modritz for the initiation of this study; None of the individuals listed were financially compensated.

## Funding

This research was funded by the Austrian Federal Ministry for Arts, Culture, Civil Service, and Sport, grant number GZ: 2022-0.865.617.

## Author information

### Author Contributions

Conceptualization, G.J. and M.N.M.v.P.; methodology, G.J.; formal analysis, G.J.; investigation, G.J.; resources, G.J.; data curation, G.J.; writing—original draft preparation, G.J.; writing—review and editing, G.J., R.K., and M.N.M.v.P.; visualization, G.J.; supervision, M.N.M.v.P.; project administration, G.J.; funding acquisition, G.J. All authors read and agreed to the published version of the manuscript.

### Corresponding author

Correspondence to Gerald Jarnig.

### Ethics declarations

#### Ethical approval and consent to participate

This study was conducted according to the guidelines of the Declaration of Helsinki and approved by the Research Ethics Committee of the University of Graz, Styria, Austria (GZ. 39/23/63 ex 2018/19). All participants gave their consent to participate in the study.

#### Consent for publication

Not applicable.

#### Competing interests

The authors declare no conflicts of interest. The funders had no role in the design of the study; in the collection, analyses, or interpretation of data; in the writing of the manuscript, or in the decision to publish the results.

## Notes

### Competing Interest Statement

The authors have declared no competing interest.

### Clinical Trial

Trial registration
DRKS00025515

