## supplements Jarnig et al for "Effects of an innovative daily physical activity intervention on the health and fitness status of primary school children: a cluster randomized controlled trial"

**Sampling calculation and school randomization**

The sample size was calculated using the G*Power calculation tool [1], which is available free of charge on the Internet.

The following Parameters were set for the calculation:

Tails: two; Effect size d: 0.30; Alpha error: 0.05; Power: 0.80; Allocation N1/N2: 1

Using these parameters, the sample calculation resulted in a total of 352 children (176 children each in the intervention and control group).

Expecting a 20% dropout rate, a sample size of minimum 423 children had to be recruited.

Based on the school and child numbers from the 2019/20 school year, the required percentage distribution of children participating in the study was as follows:

Number of children: (school year 2019/20 - fourth grade primary school)

Klagenfurt urban district = 894 (62.1%)

Klagenfurt rural district = 546 (37.9%)

Total number of schoolchildren = 1440 (100%)

Based on this calculation (about 60% of the schoolchildren were registered in the urban district), it was decided that 12 schools or at least 22 school classes would be invited to participate in the study. Seven were invited from the district of Klagenfurt–city and five from the district of Klagenfurt–country. The selected primary schools were divided into intervention and control groups, using a computerized random number generator, stratified for rural and urban districts.

**Table S1.** Raw fitness data and standard deviation scores (SDS) at baseline and follow-up by intervention group (intervention vs control).

| **health and fitness data** | **Variable** |  | |  | |
| --- | --- | --- | --- | --- | --- |
|  |  | **IG (*n*=228)** | | **CG (*n*=184)** | |
|  |  | **Baseline** | **Follow-up** | **Baseline** | **Follow-up** |
| **Anthropometrics** | **BMI, mean (SD) in kg/m^2^** | 17.84 (3.09) | 18.29 (3.29) | 18.35 (3.52) | 18.82 (3.83) |
|  | **BMI_IOTF_ SDS, mean (SD)** | 0.45 (1.03) | 0.44 (1.06) | 0.59 (1.05) | 0.57 (1.08) |
|  | **WtHR, mean (SD)** | 0.448 (0.061) | 0.438 (0.057) | 0.439 (0.059) | 0.445 (0.067) |
|  | **WtHR SDS, mean (SD)** | -0.10 (1.22) | 0.04 (1.17) | 0.08 (1.23) | -0.08 (1.29) |
| **Fitness** | **6MR, mean (SD) in m** | 858 (177) | 952 (167) | 827 (161) | 905 (174) |
|  | **SLJ, mean (SD) in cm** | 138 (20) | 146 (21) | 138 (20) | 144 (21) |
|  | **MB2kg, mean (SD) in cm** | 328 (53) | 359 (61) | 327 (52) | 365 (60) |
|  | **JS, mean (SD) in No.** | 32.0 (5.8) | 34.9 (5.8) | 30.6 (6.1) | 33.1 (5.5) |
|  | **PU, mean (SD) in No.** | 11.7 (3.5) | 13.3 (3.6) | 12.0 (3.6) | 14.0 (3.9) |
|  | **VSR, mean (SD) in cm** | 15.4 (8.5) | 15.6 (9.9) | 17.3 (8.9) | 15.9 (9.2) |
|  | **4 × 10SHR, mean (SD) in sec.** | 13.42 (1.11) | 12.84 (0.97) | 13.54 (1.23) | 13.05 (1.19) |
|  | **6MR SDS, mean (SD)** | -0.41 (1.26) | 0.05 (1.19) | -0.63 (1.15) | -0.28 (1.17) |
|  | **SLJ SDS, mean (SD)** | 0.23 (1.04) | 0.34 (1.00) | 0.24 (1.00) | 0.25 (1.00) |
|  | **MB2kg SDS, mean (SD)** | -0.07 (0.96) | -0.10 (1.05) | -0.09 (0.94) | -0.05 (1.05) |
|  | **JS SDS, mean (SD)** | 0.30 (0.81) | 0.43 (0.78) | 0.07 (0.85) | 0.17 (0.77) |
|  | **PU z-value, mean (SD)** | 0.22 (1.03) | 0.60 (1.07) | 0.32 (1.06) | 0.77 (1.14) |
|  | **VSR SDS, mean (SD)** | 0.08 (1.20) | 0.12 (1.31) | 0.34 (1.22) | 0.14 (1.25) |
|  | **4 × 10SHR SDS, mean (SD)** | 0.03 (0.92) | 0.26 (0.88) | -0.08 (0.98) | 0.09 (1.05) |

Data are the *n* or mean (SD). IG = Intervention Group, CG = Control Group; *n* = number of participations, BMI = body mass index, IOTF = International Obesity Taskforce; WtHR = waist-to-height ratio; 6MR = six-minute run; SLJ = standing long jump; MB2kg = medicine ball throw (2 kg); JS = jumping sideways; PU = push up; VSR = V sit-and-reach test; 4×10SHR = 4 × 10 m shuttle run; SD = standard deviation; m = meter; cm = centimeter; s = seconds, kg = kilogram, No = Number, SDS = standard deviation score; *z*-value = traditional *z*-score standardization.

**Table S2.** Raw fitness data and standard deviation scores (SDS) at baseline and follow-up, by sports club membership (yes or no) and intervention group (intervention vs control).

| **health and fitness data** | **Variable** | **No Sports Club** | | | | **Sports Club** | | | |
| --- | --- | --- | --- | --- | --- | --- | --- | --- | --- |
|  |  | **IG *(n*=127)** | | **CG (*n*=114)** | | **IG (*n*=101)** | | **CG (*n*=70)** | |
|  |  | **Baseline** | **Follow-up** | **Baseline** | **Follow-up** | **Baseline** | **Follow-up** | **Baseline** | **Follow-up** |
| **Anthropometrics** | **BMI, mean (SD) in kg/m^2^** | 17.97 (3.38) | 18.44 (3.52) | 18.74 (3.75) | 19.25 (4.14) | 17.68 (2.70) | 18.09 (2.98) | 17.72 (3.05) | 18.13 (3.17) |
|  | **BMI_IOTF_ SDS, mean (SD)** | 0.47 (1.09) | 0.47 (1.12) | 0.70 (1.08) | 0.68 (1.12) | 0.43 (0.95) | 0.41 (0.98) | 0.42 (0.99) | 0.40 (1.00) |
|  | **WtHR, mean (SD)** | 0.453 (0.065) | 0.441 (0.062) | 0.443 (0.064) | 0.448 (0.073) | 0.442 (0.056) | 0.433 (0.051) | 0.431 (0.049) | 0.440 (0.057) |
|  | **WtHR SDS, mean (SD)** | -0.23 (1.30) | -0.03 (1.28) | -0.03 (1.32) | -0.15 (1.36) | 0.05 (1.11) | 0.13 (1.03) | 0.25 (1.06) | 0.03 (1.19) |
| **Fitness** | **6MR, mean (SD) in m** | 825 (164) | 932 (156) | 786 (142) | 854 (157) | 898 (184) | 978 (178) | 894 (170) | 987 (169) |
|  | **SLJ, mean (SD) in cm** | 135 (21) | 144 (23) | 134 (18) | 140 (20) | 140 (19) | 149 (18) | 145 (21) | 153 (21) |
|  | **MB2kg, mean (SD) in cm** | 319 (54) | 354 (65) | 319 (50) | 357 (59) | 340 (51) | 365 (56) | 340 (55) | 378 (59) |
|  | **JS, mean (SD) in No.** | 31.2 (5.8) | 33.9 (6.1) | 28.5 (5.1) | 31.2 (4.7) | 33.1 (5.8) | 36.1 (5.2) | 33.9 (6.1) | 36.2 (5.1) |
|  | **PU, mean (SD) in No.** | 11.3 (3.4) | 12.7 (3.6) | 11.3 (3.7) | 13.0 (4.0) | 12.1 (3.6) | 14.2 (3.5) | 13.3 (3.0) | 15.6 (3.0) |
|  | **VSR, mean (SD) in cm** | 16.3 (8.2) | 17.1 (9.4) | 16.7 (9.3) | 15.1 (9.6) | 14.3 (8.7) | 13.7 (10.1) | 18.2 (8.3) | 17.3 (8.5) |
|  | **4 × 10SHR, mean (SD) in sec.** | 13.59 (1.16) | 12.97 (1.04) | 13.85 (1.18) | 13.36 (1.16) | 13.22 (1.02) | 12.66 (0.84) | 13.02 (1.14) | 12.54 (1.08) |
|  | **6MR SDS, mean (SD)** | -0.58 (1.20) | -0.02 (1.15) | -0.90 (1.04) | -0.58 (1.10) | -0.19 (1.30) | 0.15 (1.25) | -0.20 (1.20) | 0.21 (1.14) |
|  | **SLJ SDS, mean (SD)** | 0.19 (1.11) | 0.29 (1.09) | 0.07 (0.94) | 0.05 (0.97) | 0.28 (0.96) | 0.41 (0.88) | 0.52 (1.03) | 0.57 (0.96) |
|  | **MB2kg SDS, mean (SD)** | -0.14 (1.03) | -0.09 (1.16) | -0.19 (0.90) | -0.15 (1.05) | 0.02 (0.93) | -0.12 (0.93) | 0.08 (0.99) | 0.10 (1.05) |
|  | **JS SDS, mean (SD)** | 0.19 (0.82) | 0.31 (0.84) | -0.22 (0.72) | -0.10 (0.67) | 0.44 (0.77) | 0.58 (0.67) | 0.55 (0.84) | 0.61 (0.72) |
|  | **PU z-value, mean (SD)** | 0.13 (1.00) | 0.41 (1.07) | 0.09 (1.09) | 0.48 (1.19) | 0.35 (1.06) | 0.83 (1.03) | 0.68 (0.90) | 1.24 (0.87) |
|  | **VSR SDS, mean (SD)** | 0.16 (1.15) | 0.26 (1.27) | 0.20 (1.27) | -0.04 (1.29) | -0.03 (1.28) | -0.06 (1.34) | 0.56 (1.09) | 0.43 (1.11) |
|  | **4 × 10SHR SDS, mean (SD)** | -0.06 (0.95) | 0.21 (0.95) | -0.32 (0.90) | -0.17 (0.96) | 0.14 (0.87) | 0.32 (0.79) | 0.32 (0.99) | 0.51 (1.04) |

Data are the *n* or mean (SD). IG = Intervention Group, CG = Control Group; *n* = number of participations, BMI = body mass index, IOTF = International Obesity Taskforce; WtHR = waist-to-height ratio; 6MR = six-minute run; SLJ = standing long jump; MB2kg = medicine ball throw (2 kg); JS = jumping sideways; PU = push up; VSR = V sit-and-reach test; 4×10SHR = 4 × 10 m shuttle run; SD = standard deviation; m = meter; cm = centimeter; s = seconds, kg = kilogram, No = Number, SDS = standard deviation score; *z*-value = traditional *z*-score standardization;
